# Temporal Validation and Simplification of Mortality Prediction for Perioperative Cardiopulmonary Resuscitation

**DOI:** 10.64898/2026.09.15.26363126

**Authors:** Lucy Chen, Samuel Justice, Siva Muthupalaniappan, Sachin J. Shah, Matthew B. Allen

## Abstract

**Background:** Perioperative cardiac arrest is associated with high mortality, but population-level estimates of outcomes after perioperative cardiopulmonary resuscitation (CPR) do not account for differences in risk among individual patients. We previously developed the CPR OutcoMes Prediction for Arrest in Surgical Settings (COMPASS) model to estimate 30-day mortality after cardiac arrest requiring CPR on the day of surgery. The model incorporates 33 preoperative predictors and has not been evaluated in a temporally distinct cohort. We therefore sought to temporally validate COMPASS and to develop and temporally validate a simplified point-based mortality score.

**Methods:** We performed a retrospective prognostic study of adults undergoing noncardiac surgery who experienced cardiac arrest requiring CPR on postoperative day 0 in the American College of Surgeons National Surgical Quality Improvement Program. The primary outcome was 30-day mortality. The published COMPASS model, developed using 2012–2023 data, was applied without refitting to the 2024 cohort. A simplified point-based score was developed in the 2012–2023 cohort and temporally validated in 2024. Performance was assessed using discrimination, calibration, and Brier score.

**Results:** The 2024 temporal validation cohort included 429 patients, of whom 274 (63.9%) died within 30 days. The COMPASS model had an area under the receiver operating characteristic curve (AUROC) of 0.82 (95% CI, 0.78–0.86), calibration intercept of 0.24, calibration slope of 1.10, and Brier score of 0.17. The simplified 3-variable score incorporating age, American Society of Anesthesiologists physical status, and case urgency had an AUROC of 0.81 (95% CI, 0.77–0.86), calibration intercept of 0.19, calibration slope of 1.25, and Brier score of 0.17.

**Conclusions:** In a temporally distinct cohort, the published COMPASS model retained predictive performance for 30-day mortality after perioperative CPR. The simplified score showed similar performance. These findings support further evaluation of both approaches as tools for individualized estimation of perioperative CPR mortality.

## INTRODUCTION

Cardiac arrest requiring cardiopulmonary resuscitation (CPR) on the day of surgery is associated with high short-term mortality.^1,2^ For patients with preexisting limitations on resuscitation, the American Society of Anesthesiologists and American College of Surgeons recommend preoperative discussion and reconsideration of how those limitations should apply during anesthesia and surgery.^3,4^ Geriatric surgery standards and Age-Friendly Health Systems similarly emphasize preoperative assessment of patients’ goals, treatment preferences, and code status.^5,6^ These discussions require context-specific prognostic information that is relevant to the individual patient rather than population-level estimates alone.

Despite growing emphasis on shared decision making regarding perioperative CPR, clinicians have lacked practical tools for estimating an individual patient’s risk of mortality.^7,8^ We previously developed the CPR OutcoMes Prediction for Arrest in Surgical Settings (COMPASS) model using ACS-NSQIP data from 2012 through 2023 to estimate 30-day mortality after cardiac arrest requiring CPR on the day of surgery.^8^ Although COMPASS demonstrated strong discrimination in internal validation, it incorporates 33 preoperative predictors and implementation requires computational support.^8^ Its performance in a temporally distinct cohort has not been established, and its complexity may limit rapid use during time-constrained perioperative discussions.

We therefore sought to temporally validate the published COMPASS mortality model in a distinct 2024 cohort and to develop and temporally validate a simplified point-based mortality score derived from COMPASS predictors. We hypothesized that the published model would retain predictive performance in the 2024 cohort and that a simplified score would preserve much of the performance of the full model.

## METHODS

### Data Source and Study Population

We performed a retrospective prognostic study using data from the American College of Surgeons National Surgical Quality Improvement Program (ACS-NSQIP), a multicenter national surgical registry based on standardized data collection by trained and certified reviewers.^9^ The temporal validation cohort included adults aged 18 years or older undergoing noncardiac surgery who experienced cardiac arrest requiring cardiopulmonary resuscitation (CPR) on the day of surgery (postoperative day 0 [POD0]) in the 2024 ACS-NSQIP dataset. Eligibility criteria mirrored those used in the original COMPASS study, which was developed using ACS-NSQIP data from 2012 through 2023.

The validation sample comprised all eligible patients in the 2024 dataset. We performed a sample-size assessment for validation of an existing multivariable prediction model using the pmvalsampsize package in R.^10,11^ Assuming a 30-day mortality rate of 60% and an area under the receiver operating characteristic curve (AUROC) of 0.80 based on the original COMPASS study,^8^ at least 315 patients were required for the lower bound of the 95% CI for the AUROC to exceed 0.75.

Cardiac arrest was defined by ACS-NSQIP as the absence of cardiac rhythm or the presence of a chaotic cardiac rhythm requiring initiation of CPR.^9^ The primary outcome was ascertained for all eligible patients, so no patients were excluded. The study was deemed exempt from institutional review board review because it used deidentified data. The study was reported in accordance with the TRIPOD+AI guideline.^12^

### Outcomes

The primary outcome was all-cause mortality within 30 days of cardiac arrest on the day of the index operation.

### Temporal Validation of the Published COMPASS Mortality Model

Weight loss and dyspnea were unavailable in the 2024 ACS-NSQIP dataset, precluding direct calculation of the revised Risk Analysis Index (RAI).^9,13^ Weight loss, dyspnea, and RAI were therefore imputed separately for all patients using the corresponding bagged imputation models developed in the historical COMPASS cohort and applied to covariates available in 2024.^8,14^ The imputation models treated each variable as continuous, allowing imputed values between observed category values. The published COMPASS model was then applied to the 2024 cohort without refitting or recalibration.

### Development of a Simplified Point-Based Mortality Score

We developed a simplified point-based mortality score using the historical 2012–2023 COMPASS development cohort. Candidate variables comprised the 33 preoperative predictors included in the published COMPASS mortality model.^8^ Score development prioritized parsimony while seeking to preserve predictive performance. The 2024 validation cohort was not used for variable selection, score construction, or optimization.

Categorical predictors were encoded using integer values, and continuous predictors were rounded to the nearest integer when applicable. Integer point coefficients were estimated using a randomized optimization algorithm implemented with the riskscores package in R.^15^ The optimization constrained candidate models to use integer coefficients and a limited number of predictors, thereby jointly optimizing parsimony and predictive performance.^15^ The final score specification was selected using cross-validation, and total point values were mapped to predicted 30-day mortality risk.

As a sensitivity analysis, we assessed whether adding predictors to the parsimonious score materially improved predictive performance. Starting with the selected score, we sequentially added one predictor at a time. At each step, the predictor selected for addition was the one most frequently chosen across 1,000 runs of the optimization algorithm. Expanded scores were compared with the selected score using discrimination, calibration, and Brier score.

### Validation of the Simplified Score

The simplified score was evaluated in the same 2024 temporal validation cohort. Observed 30-day mortality was also examined across score categories.

### Statistical Analysis

Baseline characteristics of the 2024 temporal validation cohort were summarized using counts and percentages for categorical variables and means with standard deviations or medians with interquartile ranges for continuous variables, as appropriate. For both the published COMPASS model and simplified score, discrimination was assessed using the area under the receiver operating characteristic curve (AUROC) with 95% CIs, calibration using calibration plots and calibration intercepts and slopes with 95% CIs, and overall prediction error using the Brier score.^16^ Performance of the two models was compared descriptively; no formal hypothesis tests were performed to compare model performance. Threshold-dependent measures, including accuracy, sensitivity, specificity, positive predictive value, and negative predictive value, were calculated using a predicted-mortality threshold of 0.50 and are reported in Supplemental Table 1. All analyses were performed using R version 4.6.0 (R Project for Statistical Computing).

## RESULTS

### Cohort Characteristics

The 2024 temporal validation cohort included 429 patients who experienced cardiac arrest requiring CPR on POD0. Median age was 70 years (IQR, 62–79), 230 patients (53.6%) were men, and 274 (63.9%) died within 30 days. Baseline demographic and clinical characteristics are shown in Table 1.

**Table 1.** Characteristics of the 2024 Temporal Validation Cohort.

| Variable |  | Overall<br>n = 429 | Alive<br>n = 155<br>(36.1%) | Dead<br>n = 274<br>(63.9%) | % Missing |
| --- | --- | --- | --- | --- | --- |
| n |  |  | 155 | 274 |  |
| Age | 18-49 | 45 (10.5) | 22 (14.2) | 23 (8.4) | 0.0 |
|  | 50-64 | 96 (22.4) | 43 (27.7) | 53 (19.3) |  |
|  | 65-74 | 122 (28.4) | 47 (30.3) | 75 (27.4) |  |
|  | 75-84 | 121 (28.2) | 35 (22.6) | 86 (31.4) |  |
|  | 85+ | 45 (10.5) | 8 (5.2) | 37 (13.5) |  |
| Sex | Female | 199 (46.4) | 63 (40.6) | 136 (49.6) | 0.0 |
|  | Male | 230 (53.6) | 92 (59.4) | 138 (50.4) |  |
| Race | Black | 63 (14.7) | 20 (12.9) | 43 (15.7) | 0.0 |
|  | White | 256 (59.7) | 88 (56.8) | 168 (61.3) |  |
|  | Other<sup>a</sup> | 39 (9.1) | 17 (11.0) | 22 (8.0) |  |
|  | Unknown | 71 (16.6) | 30 (19.4) | 41 (14.2) |  |
| Ethnicity | Hispanic | 37 (8.6) | 19 (12.3) | 18 (6.6) | 0.0 |
|  | Non-Hispanic | 327 (76.2) | 110 (71.0) | 217 (79.2) |  |
|  | Unknown | 65 (15.2) | 26 (16.8) | 39 (14.2) |  |
| Admitted from home | Yes | 352 (82.1) | 138 (89.0) | 214 (78.1) | 0.0 |
|  | No | 77 (17.9) | 17 (11.0) | 60 (21.9) |  |
| ASA Physical Status | 1-2 | 38 (8.9) | 34 (21.9) | 4 (1.5) | 0.7 |
|  | 3 | 168 (39.4) | 83 (53.5) | 85 (31.4) |  |
|  | 4 | 150 (35.2) | 34 (21.9) | 116 (42.8) |  |
|  | 5 | 70 (16.4) | 4 (2.6) | 66 (24.4) |  |
| Functional Status | Independent | 360 (89.3) | 138 (92.0) | 222 (87.7) | 6.1 |
|  | Partially Dependent | 33 (8.2) | 11 (7.3) | 22 (8.7) |  |
|  | Totally Dependent | 10 (2.5) | 1 (0.7) | 9 (3.6) |  |
| BMI, kg/m<sup>2</sup> mean (SD) |  | 28.54 (7.43) | 28.8 (6.2) | 28.4 (8.1) | 9.3 |
| Smoking |  | 81 (18.9) | 29 (18.7) | 52 (19.0) | 0.0 |
| Ventilator dependent |  | 50 (11.7) | 2 (1.3) | 48 (17.5) | 0.0 |
| Diabetes |  | 40 (9.3) | 15 (9.7) | 25 (9.1) | 0.0 |
| COPD |  | 39 (9.1) | 13 (8.4) | 26 (9.5) | 0.0 |
| Sepsis or septic shock | SIRS/Sepsis | 83 (19.3) | 22 (14.2) | 61 (22.3) | 0.0 |
|  | Septic Shock | 110 (25.6) | 6 (3.9) | 104 (38.0) |  |
|  | None | 236 (55.0) | 127 (81.9) | 109 (39.8) |  |
| Cancer |  | 25 (5.8) | 5 (3.2) | 20 (7.3) | 0.0 |
| Ascites |  | 31 (7.2) | 4 (2.6) | 27 (9.9) | 0.0 |
| Hypertension on medication |  | 273 (63.6) | 99 (63.9) | 174 (63.5) | 0.0 |
| Congestive heart failure |  | 83 (19.3) | 28 (18.1) | 55 (20.1) | 0.0 |
| Transfusion<sup>b</sup> |  | 49 (11.4) | 8 (5.2) | 41 (15.0) | 0.0 |
| Bleeding disorder |  | 83 (19.3) | 16 (10.3) | 67 (24.5) | 0.0 |
| Dialysis |  | 37 (8.6) | 8 (5.2) | 29 (10.6) | 0.0 |
| Acute renal failure |  | 20 (4.7) | 2 (1.3) | 18 (6.6) | 0.0 |
| Hematocrit, mean (SD), % |  | 36.44 (7.81) | 38 (7) | 36 (8) | 4.4 |
| Platelets, mean (SD), 1,000s/μL | | 251.48 (136.95) | 266 (127) | 244 (142) | 4.9 |
| White Blood Cells, mean (SD), 1,000s/μL | | 10.61 (6.72) | 8.8 (4.2) | 11.6 (7.6) | 5.6 |
| Sodium, mean (SD), mEq/L |  | 137.37 (5.03) | 138 (3) | 137 (6) | 4.9 |
| Blood Urea Nitrogen, mean (SD), mg/dL |  | 28.45 (19.20) | 22 (16) | 32 (20) | 8.6 |
| Creatinine, mean (SD), mg/dL |  | 1.73 (1.51) | 1.4 (1.6) | 1.9 (1.5) | 4.2 |
| Procedure Urgency | Elective | 179 (41.7) | 105 (67.7) | 74 (27.0) | 0.0 |
|  | Urgent | 58 (13.5) | 20 (12.9) | 38 (13.9) |  |
|  | Emergent | 192 (44.8) | 30 (19.4) | 162 (59.1) |  |
| Operative Stress Score | 1-2 | 56 (14.2) | 32 (22.5) | 24 (9.5) | 7.9 |
|  | 3 | 218 (55.2) | 77 (54.2) | 141 (55.7) |  |
|  | 4-5 | 121 (30.6) | 33 (23.2) | 88 (34.8) |  |

Data are presented as No. (%) unless otherwise indicated. Percentages for categorical variables are calculated among patients with non-missing data. Missingness is calculated relative to the full validation cohort (N = 429).
Abbreviations: ASA, American Society of Anesthesiologists; COPD, chronic obstructive pulmonary disease; SD, standard deviation.
<sup>a</sup> Other race includes American Indian or Alaska Native, Asian, Native Hawaiian or Pacific Islander, or another reported race.
<sup>b</sup> Preoperative transfusion indicates receipt of at least 1 unit of packed red blood cells or whole blood within 72 h before surgery.

### Temporal Validation of the Published COMPASS Mortality Model

When applied to the 2024 cohort, the published COMPASS mortality model had an AUROC of 0.82 (95% CI, 0.78–0.86) (Table 2). The calibration intercept was 0.24 (95% CI, 0.01–0.48), and the calibration slope was 1.10 (95% CI, 0.88–1.34) (Table 2). The Brier score was 0.17. The calibration plot showed generally close agreement between predicted and observed mortality, with modest overall underprediction (Figure 1A).

**Figure 1.**
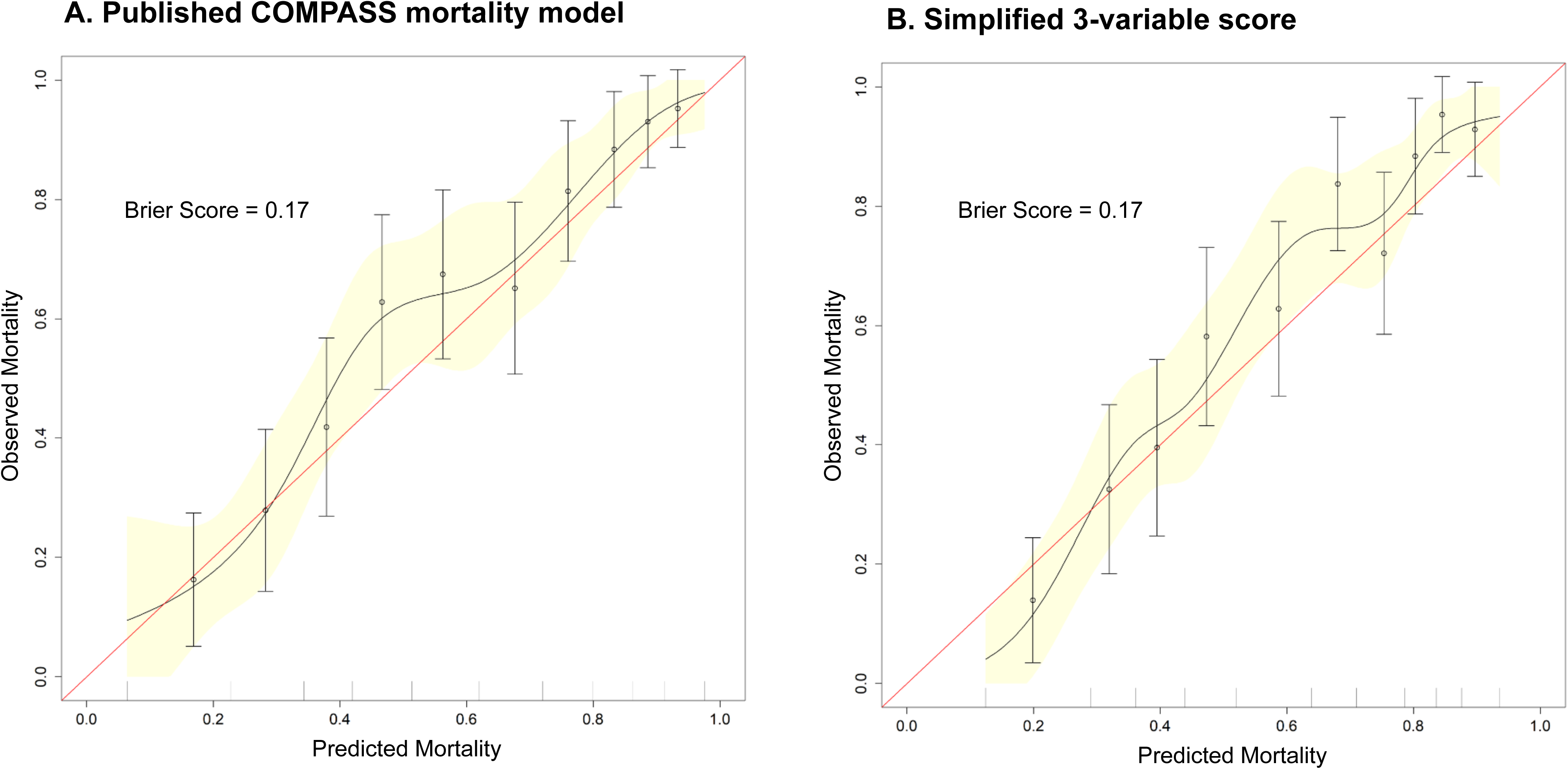
Calibration of the Published COMPASS Mortality Model and Simplified 3-Variable Score in the 2024 Temporal Validation Cohort. Points represent observed 30-day mortality within deciles of predicted risk, with vertical bars indicating 95% CIs. The solid line and shaded band represent the fitted calibration curve and its 95% CI; the dashed diagonal line represents perfect calibration. Brier scores are displayed in each panel; lower values indicate lower overall prediction error. A. Published COMPASS mortality model. B. Simplified 3-variable score.

**Table 2.** Performance of the Published COMPASS Model and Simplified Score in the 2024 Temporal Validation Cohort.

| Model | AUROC<br>(95%CI) | Calibration Intercept<br>(95% CI) | Calibration Slope<br>(95% CI) | Brier Score |
| --- | --- | --- | --- | --- |
| Published<br>COMPASS model<sup>a</sup> | 0.82 (0.78–0.86) | 0.24 (0.01–0.48) | 1.10 (0.88–1.34) | 0.17 |
| Simplified 3-variable score<sup>b</sup> | 0.81 (0.77–0.86) | 0.19 (–0.05 to 0.43) | 1.25 (1.01–1.52) | 0.17 |

Abbreviation: AUROC, area under the receiver operating characteristic curve.
Calibration intercept = 0 and calibration slope = 1 indicate ideal calibration. A positive calibration intercept indicates overall underprediction and a negative intercept indicates overall overprediction.
<sup>a</sup> Published COMPASS model applied to the 2024 cohort without refitting or recalibration.
<sup>b</sup> Simplified 3-variable score derived in the 2012–2023 COMPASS development cohort and evaluated in the 2024 temporal validation cohort.

### Development of the Simplified Point-Based Mortality Score

The simplified mortality score incorporated 3 predictors: age group, American Society of Anesthesiologists physical status, and case urgency (Table 3). For ease of interpretation, 20 points were subtracted from the original score, yielding a range of 0 to 57 points; higher scores indicated greater predicted 30-day mortality. Observed 30-day mortality increased across score categories, from 25.5% (27 of 106 patients) in the lowest category to 95.6% (65 of 68 patients) in the highest category (Figure 2).

**Figure 2.**
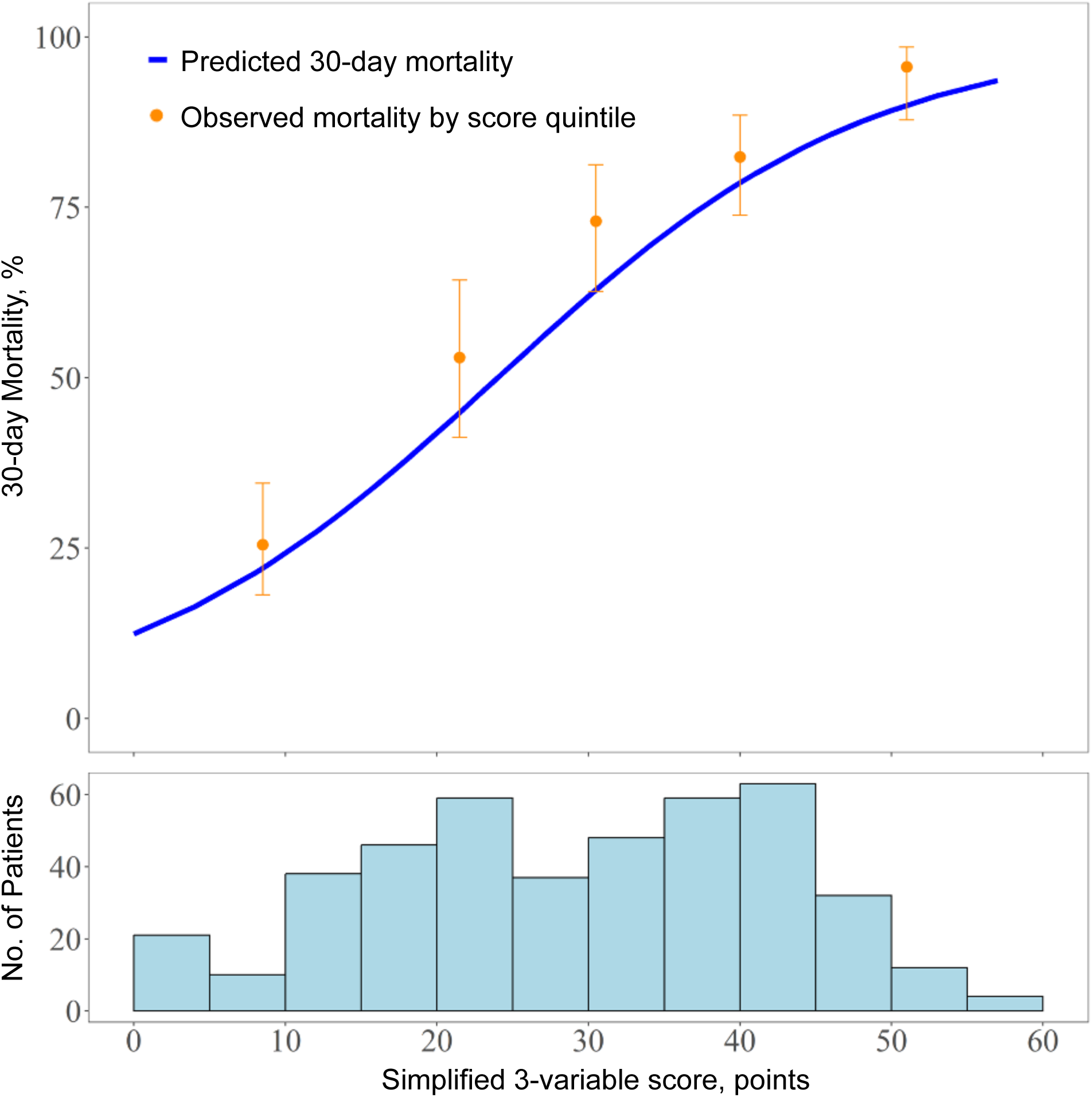
Predicted and Observed 30-Day Mortality Across the Simplified 3-Variable Score in the 2024 Temporal Validation Cohort. The solid line represents predicted 30-day mortality according to the score-to-risk function derived in the 2012–2023 COMPASS development cohort. Points represent observed 30-day mortality within score quintiles in the 2024 temporal validation cohort, with vertical bars indicating 95% CIs. The histogram shows the distribution of simplified scores in the 2024 cohort.

**Table 3.** Simplified Score for Predicting 30-Day Mortality after Perioperative Cardiac Arrest and Cardiopulmonary Resuscitation.

| Variable | Category | Points |
| --- | --- | --- |
| Age (years) | 18-49 | 4 |
|  | 50-64 | 8 |
|  | 65-74 | 12 |
|  | 75-84 | 16 |
|  | 85+ | 20 |
| ASA physical status | I-II | 9 |
|  | III | 18 |
|  | IV | 27 |
|  | V | 36 |
| Urgency | Elective | 7 |
|  | Urgent | 14 |
|  | Emergent | 21 |
|  | Total | 20-77<br>(normalize to 0-57<br>by subtracting 20) |

Add the points assigned for age, ASA physical status, and case urgency. Subtract 20 from the total to obtain the normalized score (range, 0–57); higher scores indicate greater predicted 30-day mortality.
Abbreviation: ASA, American Society of Anesthesiologists

### Validation of the Simplified Score in the 2024 Cohort

In the 2024 validation cohort, the simplified point-based mortality score had an AUROC of 0.81 (95% CI, 0.77–0.86) (Table 2). The calibration intercept was 0.19 (95% CI, −0.05 to 0.43), the calibration slope was 1.25 (95% CI, 1.01–1.52), and the Brier score was 0.17 (Table 2; Figure 1B). These measures were numerically similar to those of the published COMPASS mortality model. In sensitivity analyses, adding disseminated cancer and then operative stress score produced AUROCs of 0.82 for both expanded scores, while the Brier score remained 0.17 (Supplemental Table 2).

## DISCUSSION

In this temporal validation study, the published COMPASS mortality model retained predictive performance in a distinct 2024 cohort of patients who experienced perioperative cardiac arrest requiring CPR. A simplified point-based score incorporating age group, American Society of Anesthesiologists physical status, and case urgency demonstrated numerically similar discrimination and overall prediction error. Together, these findings support the temporal validity of individualized mortality prediction after perioperative CPR and show that much of the predictive performance of the full model can be preserved in a 3-variable point-based score.

Prior work has largely characterized outcomes after perioperative CPR using population-level estimates. In a systematic review of perioperative CPR outcomes, 24-hour survival ranged from 32.0% to 55.7%.^1^ Although such estimates describe overall prognosis, they provide limited guidance for individual patients whose risk may vary substantially according to baseline health and operative context.^2,8^ COMPASS extends this evidence base by providing individualized preoperative estimates of 30-day mortality after perioperative cardiac arrest requiring CPR.^8^ More recently, a single-center point-based score was developed to predict 24-hour survival after perioperative CPR using peri-arrest characteristics, including CPR duration and end-tidal carbon dioxide; unlike COMPASS, such variables are unavailable for preoperative prognostication.^17^

Growing emphasis on aligning perioperative care with patients’ goals and priorities creates a practical need for individualized prognostic information that can be incorporated into time-constrained clinical encounters.^5,6,18,19^ The full COMPASS model could support automated risk estimation within electronic clinical systems, whereas the simplified 3-variable score may offer a lower-complexity option when electronic decision support is unavailable. These estimates are intended to inform, rather than determine, decisions regarding perioperative CPR and should be considered alongside patients’ goals, treatment preferences, and views regarding acceptable outcomes.^19,20^ Prospective studies are needed to evaluate their integration into perioperative workflows and effects on communication and decision quality.

This study has several limitations. First, both the development and temporal validation cohorts were derived from ACS-NSQIP. The findings therefore support temporal validity within the same registry but do not establish transportability to other health systems, registries, or clinical settings.^16^ Second, weight loss and dyspnea were unavailable in the 2024 dataset, precluding direct calculation of the RAI; all three variables were therefore imputed before application of the published COMPASS model.^9,13^ This may have influenced estimates of full-model performance, although none of these variables was included in the simplified score. Third, ACS-NSQIP does not capture arrest and resuscitation characteristics such as arrest location, etiology, or CPR duration, which may further inform post-arrest prognosis.^9^ However, these factors are not available preoperatively and therefore would not contribute to the intended use of COMPASS for preoperative risk estimation. Finally, ACS-NSQIP provides 30-day outcomes and does not permit assessment of longer-term survival, functional status, or other patient-centered outcomes that may also be relevant to decisions about CPR.^21^

The published COMPASS mortality model retained predictive performance for 30-day mortality after perioperative CPR in a temporally distinct cohort, and a simplified 3-variable score retained much of that performance with substantially fewer predictors. These findings support further evaluation of both approaches for individualized mortality estimation and integration into perioperative discussions regarding CPR.

## Data Availability

Data will not be made publicly available, as it is under the governance of the American College of Surgeons' National Surgical Quality Improvement Program.

## Acknowledgments

The American College of Surgeons National Surgical Quality Improvement Program and the hospitals participating in the ACS NSQIP are the source of the data used herein; they have not verified and are not responsible for the statistical validity of the data analysis or the conclusions derived by the authors. Artificial Intelligence Use: ChatGPT (OpenAI) was used solely to assist with proofreading and checking adherence to journal administrative and formatting requirements. All resulting changes were reviewed and approved by the authors.

## Summary statement

Not applicable

## Abbreviations

ACS-NSQIP: American College of Surgeons National Surgical Quality Improvement Program;
ASA: American Society of Anesthesiologists;
AUROC: area under the receiver operating characteristic curve;
CPR: cardiopulmonary resuscitation;
COMPASS: CPR OutcoMes Prediction for Arrest in Surgical Settings;
POD: postoperative day;
RAI: Risk Analysis Index.

