## Supplemental Table for "Temporal Validation and Simplification of Mortality Prediction for Perioperative Cardiopulmonary Resuscitation"

| **Supplemental Table 1**. Threshold-Dependent Performance of the Published COMPASS Model and Simplified Score in the 2024 Temporal Validation Cohort | | | | | |
| --- | --- | --- | --- | --- | --- |
| Model | Accuracy  (95% CI) | Sensitivity  (95% CI) | Specificity (95% CI) | PPV  (95% CI) | NPV  (95% CI) |
| Published COMPASS model | 0.75  (0.71, 0.79) | 0.79  (0.74, 0.84) | 0.68  (0.60, 0.76) | 0.82  (0.76, 0.86) | 0.65  (0.57, 0.72) |
| Simplified 3-variable score | 0.76  (0.72, 0.80) | 0.80  (0.74, 0.84) | 0.70  (0.62, 0.77) | 0.82  (0.77, 0.87) | 0.66  (0.58, 0.73) |
| Classification metrics were calculated using a predicted 30-day mortality threshold of 0.50 for both models. PPV indicates positive predictive value; NPV, negative predictive value. | | | | | |

| **Supplemental Table 2. Sensitivity Analysis of Simplified Scores with Additional Predictors** | | | | |
| --- | --- | --- | --- | --- |
| Model | AUROC  (95% CI) | Calibration Intercept  (95% CI) | Calibration Slope  (95% CI) | Brier Score |
| 3 variables: age, ASA physical status, case urgency | 0.81 (0.77–0.86) | 0.19 (−0.05 to 0.43) | 1.25 (1.01–1.52) | 0.17 |
| 4 variables: + disseminated cancer | 0.82 (0.78–0.86) | 0.19 (−0.05 to 0.43) | 1.26 (1.02–1.52) | 0.17 |
| 5 variables: + disseminated cancer, Operative Stress Score | 0.82 (0.77–0.86) | 0.19 (−0.05 to 0.43) | 1.20 (0.97–1.46) | 0.17 |
| The 4- and 5-variable scores were constructed sequentially from the selected 3-variable score. At each step, the additional predictor was the one most frequently selected across 1,000 runs of the optimization algorithm. AUROC indicates area under the receiver operating characteristic curve; ASA, American Society of Anesthesiologists. | | | | |
